# Simple Words over Rich Imaging: Accurate Brain Disease Classification via Language Model Analysis of Radiological Reports

**DOI:** 10.1101/2024.11.13.24317214

**Authors:** Xin Gao, Meihui Zhang, Longfei Chen, Jun Qiu, Shanbo Zhao, Junjie Li, Tiantian Hua, Ying Jin, Zhiqiang Wu, Haotian Hou, Yunling Wang, Wei Zhao, Yuxin Li, Yunyun Duan, Chuyang Ye, Yaou Liu

## Abstract

Brain diseases exert profound detrimental effects on human health by affecting the central nervous system. Accurate automated diagnosis of brain diseases is imperative to delay the progression of illness and enhance long-term prognosis. However, existing image-based diagnostic approaches struggle to achieve satisfactory performance due to the high dimensionality of imaging data. Radiological reports, which are required in clinical routine to describe image findings, provide a more straightforward comprehension of the imaging data, yet they have been neglected in automated brain disease classification. In this work, we explore automated brain disease classification via radiological reports and language models and compare the results with conventional image-based methods. Specifically, in the report-based diagnostic approach, we fine-tune Pre-trained Language Models (PLMs) and Large Language Models (LLMs) based on the findings part of radiological reports to achieve disease classification. Four clinically relevant brain disease classification tasks were performed in our experiments, involving 12 datasets with a total number of 14,970 patients, including two independent validation sets. The best language model reached an average area under the receiver operating characteristic curve (AUC) of 84.75%, an average accuracy (ACC) of 79.48%, and an average F1-score of 79.45%. Compared with the best image-based model, it achieved an average improvement of 10.34%, 10.75%, and 9.95% in terms of AUC, ACC, and F1-score, respectively. The language model also outperformed junior radiologists by 9.47% in terms of ACC. Moreover, the report-based model exhibited better adaptability to missing image contrasts and cross-site data variability than image-based models. Together, these results show that brain disease classification via language model analysis of radiological reports can be more reliable than image-based classification, and our work demonstrates the potential of using radiological reports for accurate diagnosis of brain diseases.

## Introduction

Brain diseases seriously threaten the health and wellness of millions of people worldwide^1^. Accurate diagnosis of brain diseases, such as genetic marker testing, pathological examination, and tumor grading, enables the formulation of personalized treatment plans, which can facilitate precise early interventions^2^. To assist physicians in further improving diagnostic and treatment efficiency, various automated diagnostic models based on deep learning (DL) are developed according to the demands of different scenarios, for example, towards providing timely diagnosis in emergencies^3^ or precise diagnosis in routine clinical settings^4^.

Imaging examinations are pivotal tools for assessing the conditions of brain diseases. Currently, physicians typically scrutinize imaging data, such as 3D magnetic resonance imaging (MRI), to formulate diagnosis of brain diseases. Therefore, existing automated brain disease diagnostic methods largely rely on imaging information^5^. However, conventional image-based models struggle to produce satisfactory results. In particular, brain images are typically high-dimensional with 3D imaging consisting of multiple slices and contrasts, and they are thus complex to analyze^6,7^. It is challenging for image-based brain disease classification models to learn a sufficiently disease-relevant image representation, especially when key lesions are relatively small^8,9^. Moreover, optimal performance of an image-based model is restricted to a specific combination of input image contrasts, and the performance is also sensitive to cross-site domain shift when training and test data originate from different sites^1,10^. This compromises the applicability of image-based models in real-world scenarios, as there can be different choices of image acquisitions and data acquired at various hospitals. Therefore, it is imperative to develop innovative automated brain disease classification methods to achieve better diagnosis.

Radiological reports, which always accompany radiological images as required by clinical routine, are written by radiologists as part of their workflow. The reports comprise concise distillation of crucial information extracted by radiologists from the images and can potentially be effective for diagnosis with their reduced dimensionality^11,12^. However, brain disease classification based on radiological reports is still unexplored by existing works.

In this work, we investigated a more effective approach to brain disease classification by analyzing radiological reports with language models. In particular, as existing research has demonstrated that Pre-trained Language Models (PLMs) and Large Language Models (LLMs) have the capacity for understanding and analyzing textual information through pre-training on enormous text corpora^13–18^, both of them were adopted to take reports as input and output the disease classification results after task-specific fine-tuning. We performed experiments on four different clinically relevant brain disease classification tasks, which involved 12 datasets and a total number of 14,970 patients. The report-based model achieved excellent performance. Our best performing language model exhibited an average area under the receiver operating characteristic curve (AUC) of 84.75%, an average accuracy (ACC) of 79.48%, and an average F1-score of 79.45%. It achieved an average improvement of 10.34%, 10.75%, and 9.95% in terms of AUC, ACC, and F1-score, respectively, compared with the best image-based model, and outperformed junior radiologists by 9.47% and 9.49% in terms of ACC and F1-score, respectively. Furthermore, the results on test data without full image contrasts and external datasets show that the language model can better address missing image contrasts and cross-site data variability than image-based models. Our exploration yields a promisingly more effective strategy for accurate brain disease diagnosis in real-world clinical scenarios.

## Results

### Study design

An overview of the study design is shown in Fig. 1. We used 12 in-house datasets for the experiments, as existing public datasets for brain diseases (such as ADNI^19^ and BraTS^20^) did not contain radiological reports and public datasets with paired images and reports (such as MIMIC-CXR^21^, IU X-Ray^22^, and CheXpert^23^) focused on 2D chest imaging^23^. A detailed description of the 12 datasets are given in Table 1. Four brain disease classification tasks were considered, which were isocitrate dehydrogenase (IDH) genotyping, 1p/19q co-deletion identification, World Health Organization (WHO) grading, and brain tumor classification.

**Table 1.**
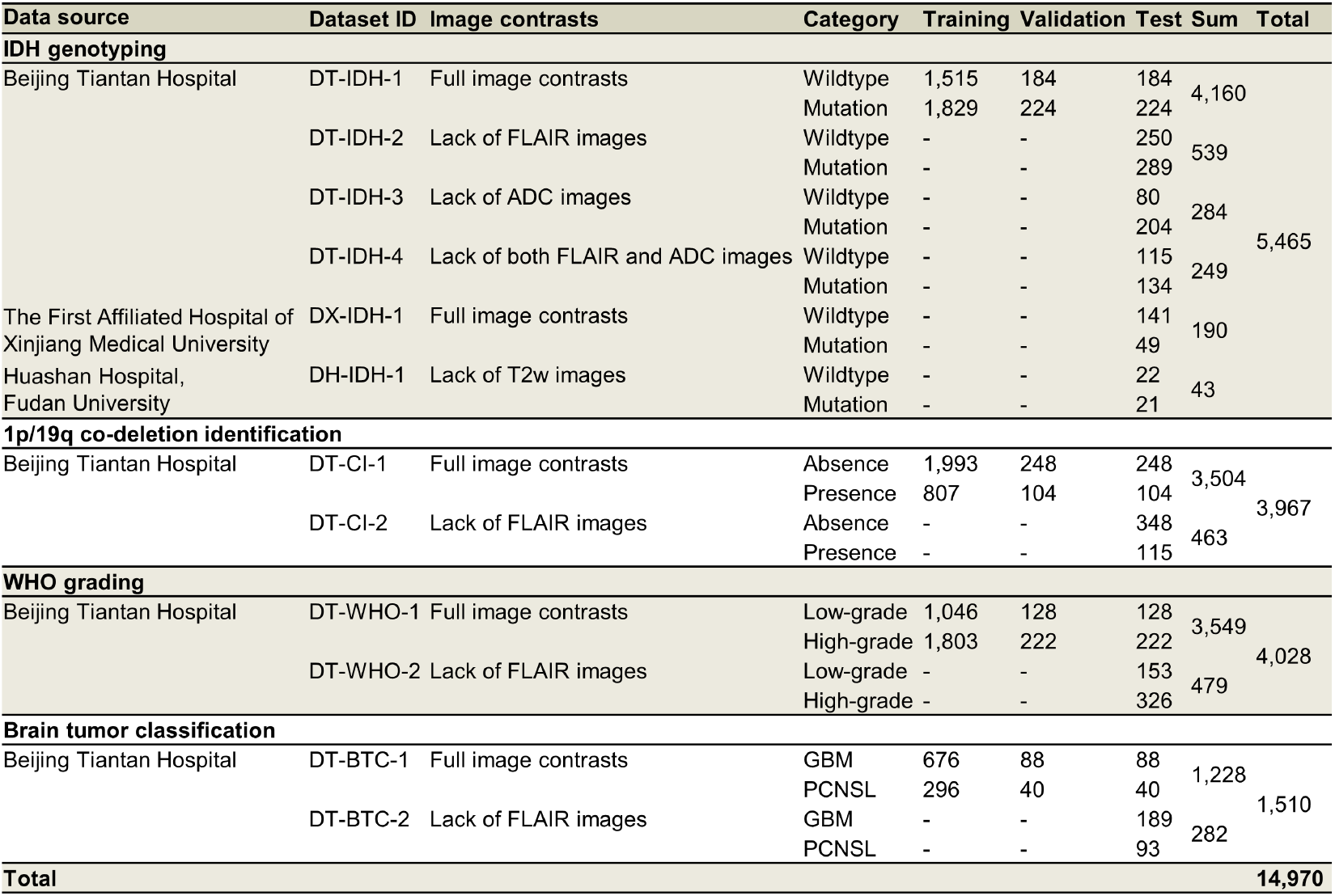
The detailed description of the in-house datasets. The numbers of subjects are indicated for each dataset, each category, and the training/validation/test split. GBM: glioblastoma; PCNSL: primary central nervous system lymphoma.

**Figure 1.**
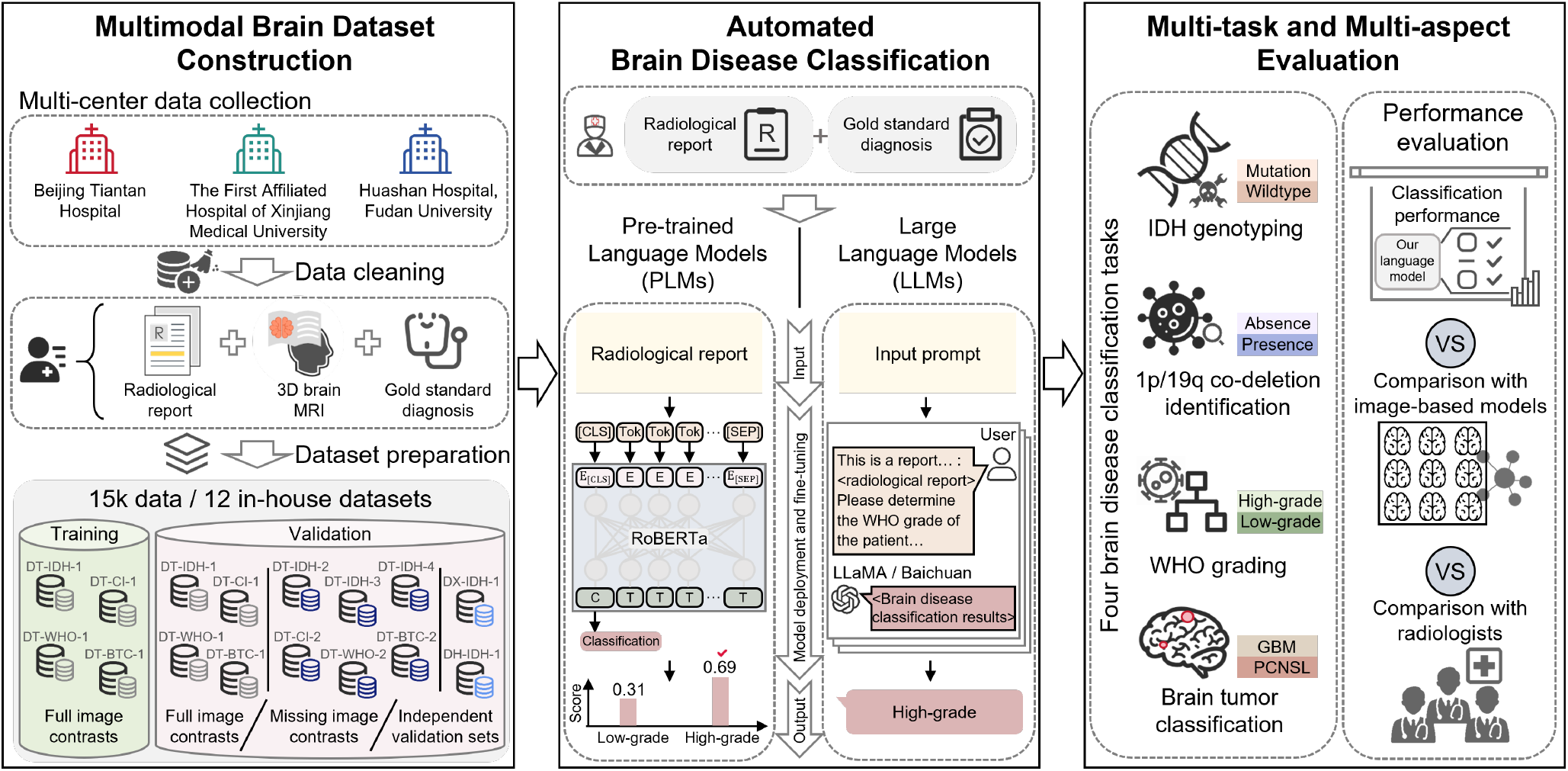
An overview of the proposed work. 12 in-house datasets were collected from three hospitals. Each dataset comprises the radiological report, 3D MRI scans, and gold standard diagnosis result for each patient. Automated brain disease classification was achieved by analyzing radiological reports with language models. The models were comprehensively evaluated on four tasks. The language models were compared with conventional image-based models and radiologists.

For all datasets, the gold standard diagnosis results were available and obtained by pathological analysis. For the task of IDH genotyping, patients were stratified according to their IDH mutation status types, categorized as either wildtype or mutation. For 1p/19q co-deletion identification, patients were classified based on the absence or presence of co-deletion in chromosomes 1p and 19q. For WHO grading, patients were categorized based on the degree of tumor malignancy as either low-grade or high-grade. For brain tumor classification, patients were categorized based on the type of tumor pathology: *glioblastoma* (GBM) or *primary central nervous system lymphoma* (PCNSL).

In four of the datasets (DT-IDH-1, DT-CI-1, DT-WHO-1, and DT-BTC-1), MRI scans were acquired with five different contrasts, including T1-weighted (T1w), T2-weighted (T2w), fluid-attenuated inversion recovery (FLAIR), T1 contrast-enhanced (T1c) and apparent diffusion coefficient (ADC) images, at Beijing Tiantan Hospital, and radiological reports were written by radiologists in the clinical routine for the images. These four datasets were used separately for the tasks of IDH genotyping, 1p/19q co-deletion identification, WHO grading, and brain tumor classification. Another dataset DX-IDH-1 was acquired at The First Affiliated Hospital of Xinjiang Medical University as an external dataset with all five MRI contrasts and reports, and it was concerned with IDH genotyping. Also, three datasets (DT-IDH-2, DT-IDH-3, and DT-IDH-4) were acquired at Beijing Tiantan Hospital without FLAIR, ADC, or both FLAIR and ADC images, respectively, and one dataset DH-IDH-1 was acquired at Huashan Hospital, Fudan University without T2w images. These datasets were concerned with IDH genotyping. Moreover, three datasets (DT-CI-2, DT-WHO-2, and DT-BTC-2) were acquired at Beijing Tiantan Hospital without FLAIR images, and they were separately used for 1p/19g co-deletion identification, WHO grading, and brain tumor classification. In the datasets with incomplete image contrasts, reports were written based on the available image contrasts, and these datasets were used for evaluating the classification performance when image contrasts were missing.

Two kinds of language models, including PLMs and LLMs, were employed for brain disease classification. For each kind of language model, a general domain version and a specific pre-trained version for Chinese were adopted. Specifically, the selected PLMs were RoBERTa-base^24^ and Chinese RoBERTa^25^, and the selected LLMs were LLaMA3-8B^26^ and Baichuan2-13B^27^. The PLMs were fine-tuned with the training sets of DT-IDH-1, DT-CI-1, DT-WHO-1, and DT-BTC-1 for their respective tasks, together with the use of their validation sets. The LLMs were fine-tuned with a parameter-efficient tuning method low-rank adaptation (LoRA)^28^, where all training sets of DT-IDH-1, DT-CI-1, DT-WHO-1, and DT-BTC-1 were combined.

For comparison, six image-based classification models were also applied. These methods included DeepRisk^29^ and 3D MedMNIST^30^, which took the MRI scans or a subset of slices as input and output the classification result, as well as 2D MedMNIST^30^, DenseNet^31^, ViT^32^, and Swin Trasnformer^33^, which first segmented the brain tumor and then made classification based on the image and segmentation results. The detailed description of the image-based models is given in “Competing image-based models”. In addition, three radiologists were invited to perform manual classification of brain diseases. The three radiologists had three, three, and ten years of clinical experience, respectively.

To evaluate the classification performance, the following metrics were computed: area under the receiver operating characteristic curve (AUC), accuracy (ACC), F1-score, sensitivity (SEN), specificity (SPEC), positive predictive value (PPV), and negative predictive value (NPV). To investigate the impact of random factors, the mean and *standard deviation* (std) of the classification performance were computed based on multiple independent repeated runs. Each run was associated with a different random seed in model training. For each task, five repeated experiments were performed. In addition, based on the results of each repeated run, Student’s *t*-tests were performed to compare the AUC and ACC between report-based and image-based models.

### The language model outperformed conventional image-based models for brain disease classification across all evaluation tasks

The language models and conventional image-based models first predicted the disease classification based on all five image contrasts for the four tasks using DT-IDH-1, DT-CI-1, DT-WHO-1, and DT-BTC-1. The comparison of the classification performance between the language model Chinese RoBERTa and conventional image-based models is summarized in Table 2. Chinese RoBERTa is shown here as it has achieved the best average performance among the language models (see “Comparison and analysis of different language models” and Extended Table E2 for the detailed comparison between language models). The fine-tuned language model consistently achieved better performance in all cases, with an average AUC of 0.848, ACC of 0.795, and F1-score of 0.795 across the four tasks. Compared with the average performance of the image-based models, the language model achieved noticeable improvements, with increases in AUC of 16.72%, 18.97%, 21.98%, and 32.27%, and in ACC of 18.47%, 15.04%, 19.58%, and 22.41% for the four tasks, respectively. In addition, compared with Swin Transformer, the best image-based model in three of four tasks in terms of AUC, our language model yielded an average ACC improvement of 10.75%, and an average AUC improvement of 10.34% across the four tasks. These results demonstrate the feasibility of using report-based language models for accurate classification of brain diseases, as well as its superiority over image-based models, which are currently the common practice.

**Table 2.**
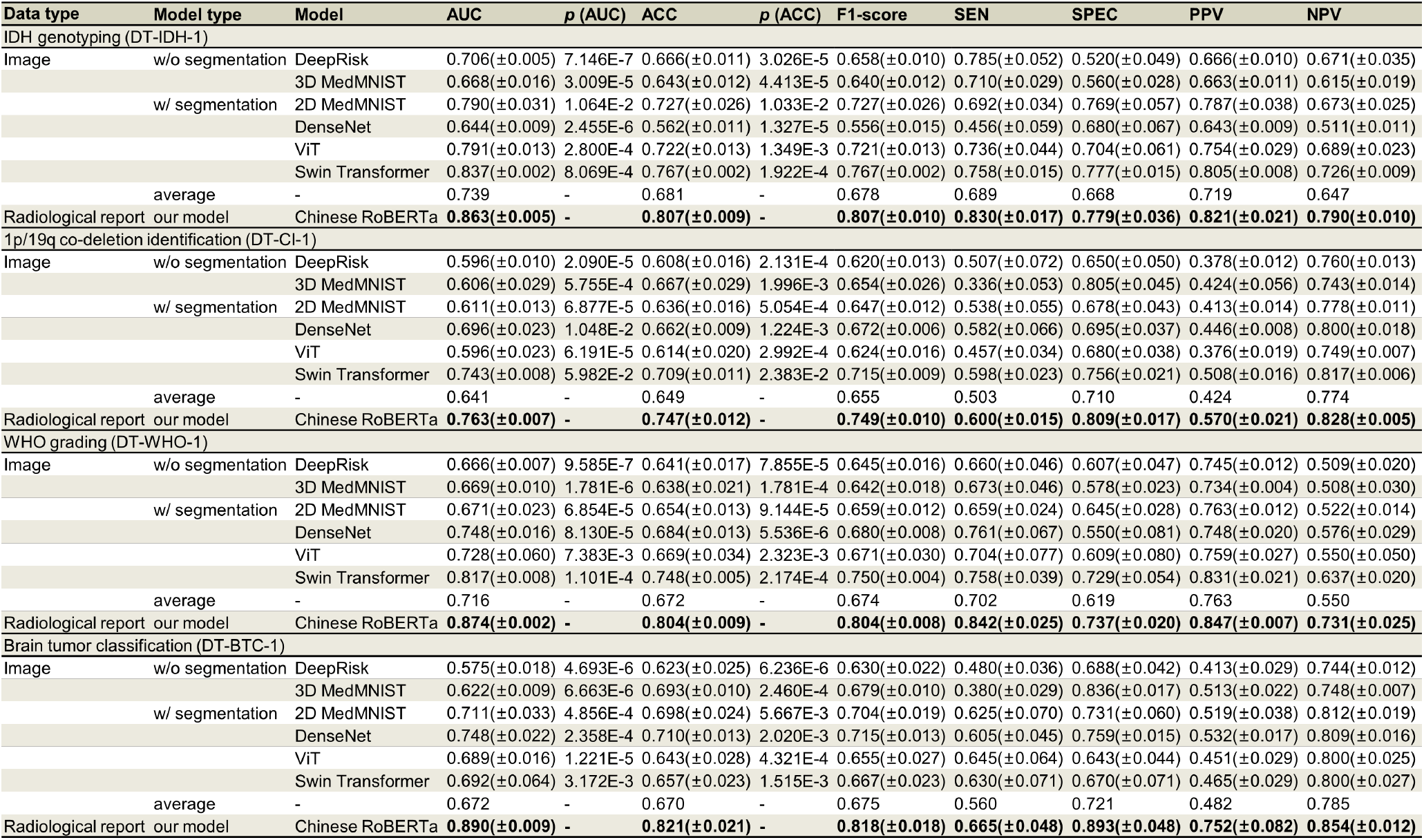
Comparison of our fine-tuned language model Chinese RoBERTa with conventional image-based models across four different brain disease classification tasks using DT-IDH-1, DT-CI-1, DT-WHO-1, and DT-BTC-1. The best result is highlighted in bold. The means and stds are presented in the format of mean(±std). *p*-values are computed with Student’s *t*-tests to compare the AUC and ACC between Chinese RoBERTa and the image-based models.

### The language model achieved better accuracy than junior radiologists

We selected the task of brain tumor classification to compare the classification performance of our fine-tuned models with radiologists, as the radiologists found the task more dependent on the visual cues from imaging and were more confident to make direct diagnosis. The comparison was based on the test set of DT-BTC-1. The radiologists were allowed to simultaneously view all five image contrasts and the corresponding radiological reports for diagnosis. The performance of the three radiologists was compared with the receiver operating characteristic (ROC) curve of Chinese RoBERTa in Figure 2a. The ACC of the two junior radiologists with three years of experience is lower than that of Chinese RoBERTa (0.821 in Table 2), and the ROC curve of Chinese RoBERTa is above the junior radiologists. These observations indicate that the report-based model outperformed the junior radiologists. Note that the ACC of the junior radiologists is higher than that of all image-based models (see Table 2). The senior radiologist made more accurate diagnosis than all image-based and report-based models.

**Figure 2.**
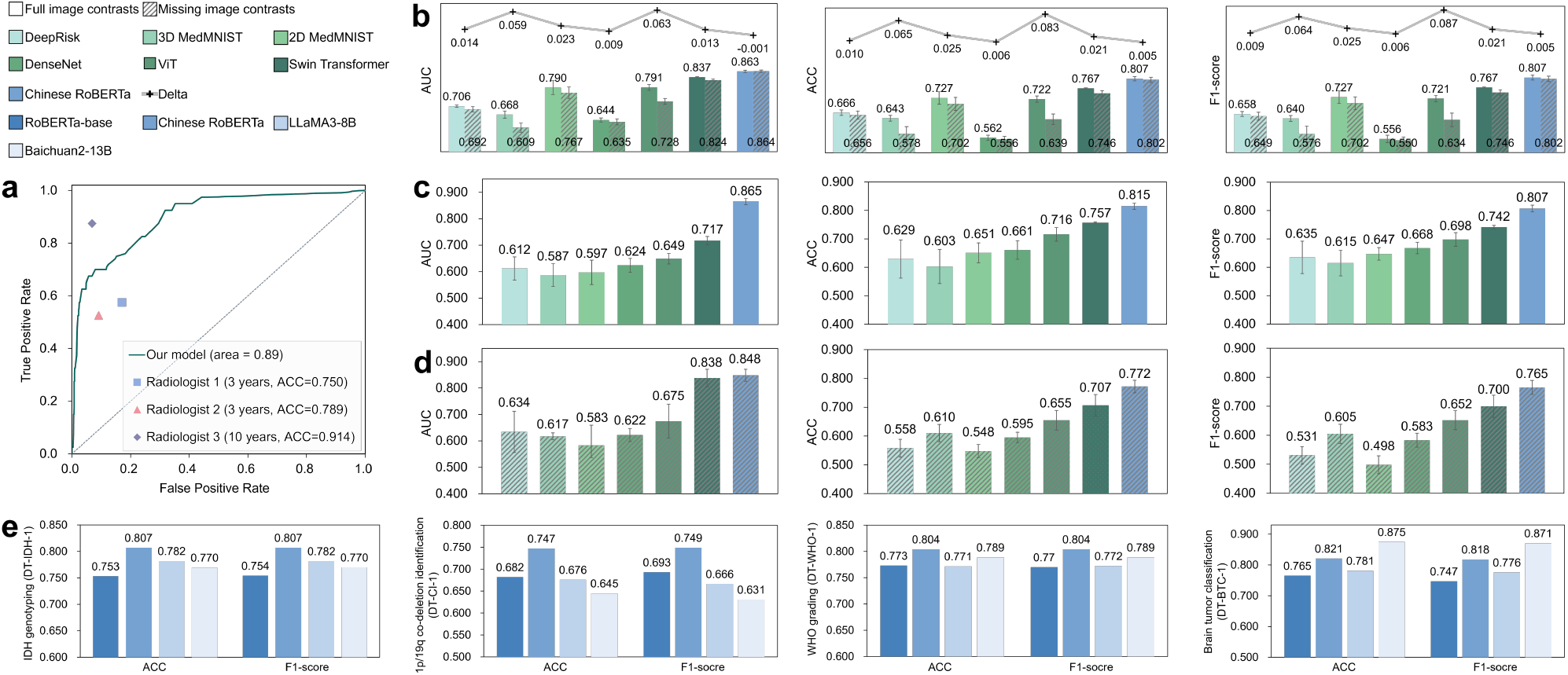
Additional comparison between Chinese RoBERTa and radiologists, image-based models, and others report-based models. **a**, The comparison of the ROC curve of Chinese RoBERTa and three radiologists. The ACC of the radiologists are also indicated. **b**, The classification performance on the simulated test set for IDH genotyping with full image contrasts (left bar) and missing FLAIR images (right bar). The reduction (Delta) in AUC, ACC, and F1-score in the case of missing contrasts is also indicated. **c** and **d**, The performance comparison between Chinese RoBERTa and image-based models for the two independent validation sets DX-IDH-1 and DH-IDH-1, respectively. **e**, The classification performance of the four language models, including RoBERTa-base, Chinese RoBERTa, LLaMA3-8B, and Baichuan2-13B, on DT-IDH-1, DT-CI-1, DT-WHO-1, and DT-BTC-1.

### The language model better handled missing image contrasts than the image-based models

Because of the different clinical conditions of patients and different levels of expertise among radiologists, it is common that for some patients not all image contrasts are acquired, which often poses challenges for conventional image-based models^34^. Therefore, we further evaluated the classification performance of the report-based and image-based models with DT-IDH-2, DT-IDH-3, DT-IDH-4, DT-CI-2, DT-WHO-2, and DT-BTC-2 in this case of missing image contrasts. The report-based Chinese RoBERTa model trained previously with full image contrasts was directly applied to the test data with missing contrasts. To apply the image-based model trained previously with full image contrasts, we synthesized the missing contrast based on acquired ones using the pGAN model^35^, and then the acquired and synthesized contrasts were jointly fed into the image-based model. The details about the pGAN model are described in “Datasets preparation and pre-processing”.

The classification performance is presented in Table 3, where FLAIR images were missing in DT-IDH-2, DT-CI-2, DT-WHO-2, and DT-BTC-2, ADC images were missing in DT-IDH-3, and both ADC and FLAIR images were missing in DT-IDH-4. Our fine-tuned language model Chinese RoBERTa outperformed six image-based models for all metrics and datasets. In particular, it achieved an average AUC of 0.792, ACC of 0.745, and F1-score of 0.747 for the six datasets. Compared with the best image-based model Swin Transformer, the average improvements in AUC, ACC, and F1-score of the language model were 14.71%, 12.07%, and 11.55%, respectively. Compared with the average performance of all image-based models, for each individual dataset, the improvements in AUC were 21.96%, 18.34%, 31.35%, 25.47%, 28.54%, and 32.08%, respectively; the improvements in ACC were 21.64%, 10.07%, 13.94%, 23.99%, 21.67%, and 26.95%, respectively; the improvements in F1-score were 22.37%, 9.39%, 16.91%, 22.39%, 26.03%, and 26.05%, respectively. These results indicate the excellent capability of the report-based model for handling the challenging scenario of missing image contrasts.

**Table 3.**
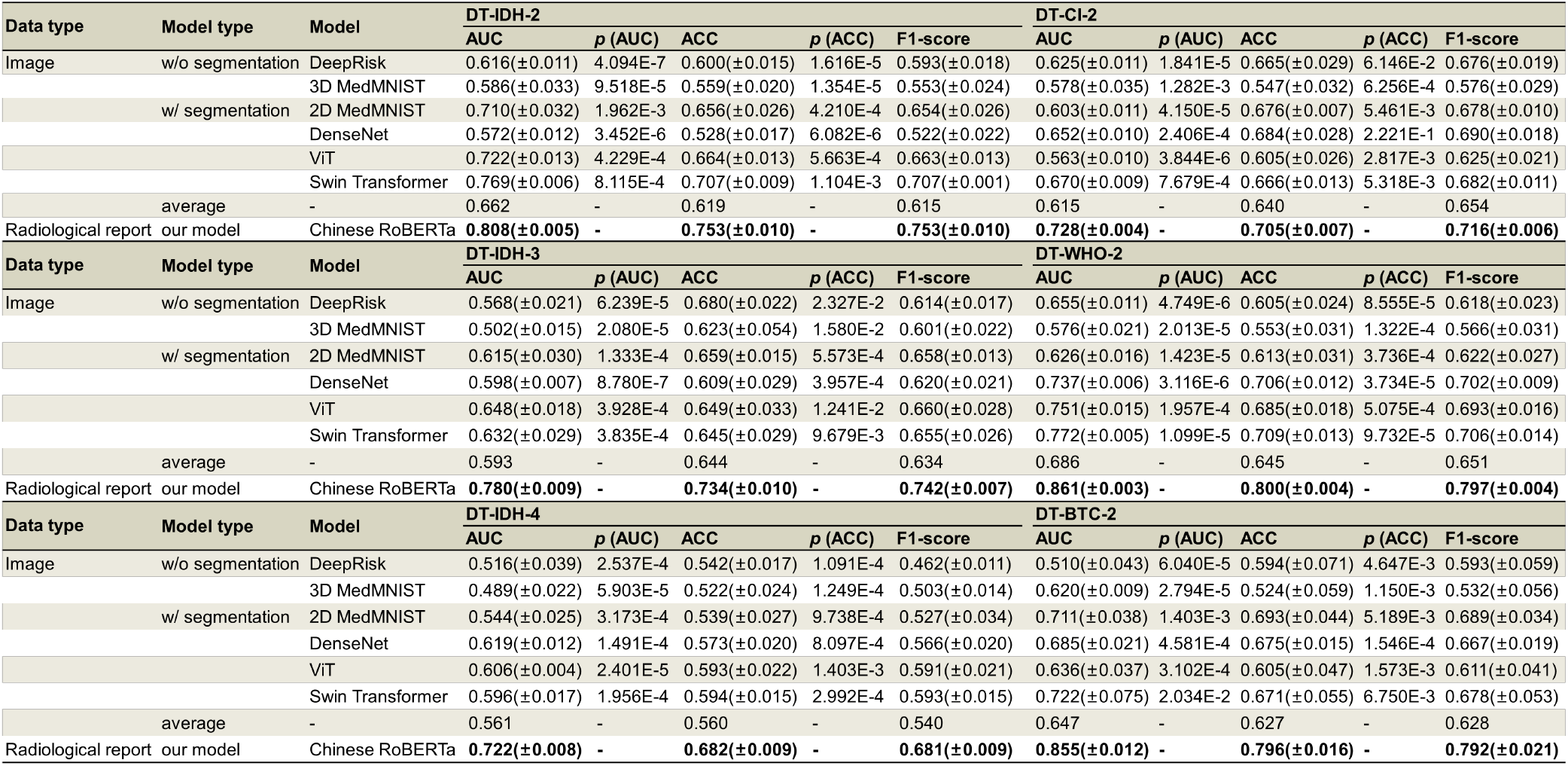
Classification performance of Chinese RoBERTa and image-based models on DT-IDH-2, DT-IDH-3, DT-IDH-4, DT-CI-2, DT-WHO-2, and DT-BTC-2 for patients with missing image contrasts. The best result is highlighted in bold.

Moreover, to further explore quantitatively the impact of missing image contrasts on the classification performance, we performed an extended experiment for the task of IDH genotyping. Specifically, we simulated a test set with missing image contrasts based on the 408 test patients with full image contrasts in DT-IDH-1. First, we removed the FLAIR images from the input of the image-based models. Second, we manually deleted the descriptions regarding FLAIR images in the radiological reports of these patients. The classification results and the reduction in AUC, ACC, and F1-score compared with the results achieved with full image contrasts are shown in Figure 2b. The performance of our language model was very close when image contrasts were missing and complete; however, the performance of the image-based models decreased noticeably with missing contrasts. These observations further show that the language model is more robust to missing image contrasts than image-based models.

### The language model better addressed cross-site data variability than image-based models

Due to the difference in the scanning device, protocol, and parameters of images acquired at different sites/hospitals, conventional image-based models may generalize poorly to an unseen site different from the training data^10^. To assess the impact of cross-site image variability on the report-based and image-based models, additional experiments were performed on the two external datasets DX-IDH-1 and DH-IDH-1. Note that in DH-IDH-1 there was also a missing image contrast, where no T2w image was available and a pre-trained pGAN model synthesized the T2w image. The results are presented in Figures 2c and 2d. The language model achieved better classification performance than the image-based models on the unseen sites. Specifically, for DX-IDH-1/DH-IDH-1 the language model achieved an improvement in AUC, ACC, and F1-score of 20.64%/1.19%, 7.66%/9.19%, 8.76%/9.28%, respectively, compared with the best image-based model Swin Transformer.

### The data pre-processing cost for the language model was lower than that of the image-based models

In addition to the benefit of better classification performance, the language model may also require less computational overhead for data pre-processing than the image-based models, as it is more convenient to pre-process texts than images. To demonstrate the advantage of the language model in terms of the pre-processing cost, we compared the time consumption for processing 3D brain imaging data versus radiological report data, and the results are shown in Extended Table E1. The image pre-processing procedures included N4 bias field correction, image co-registration, skull stripping, and optional tumor segmentation. For radiological reports, data pre-processing involved the removal of nonsensical characters (such as blank spaces, line breaks, and extraneous symbols), tokenization, padding and truncating. The pre-processing of reports required a much smaller amount of time compared to image pre-processing, which was over 6,000 times longer with tumor segmentation and 5,000 times longer without tumor segmentation. After pre-processing, the inference stage took about an average of 1.3×10^−3^ seconds per patient for the language models and 1.7×10^−3^ seconds for the image-based models. In comparison with the pre-processing time of imaging data, the inference time of both language and image-based models was considerably short and negligible. Thus, the lower time consumption of text pre-processing can allow more efficient and timely diagnosis in clinical emergency scenarios.

### Comparison and analysis of different language models

The performance of each language model for brain disease classification based on the radiological reports is summarized in Figure 2e, where the results on DT-IDH-1, DT-CI-1, DT-WHO-1, and DT-BTC-1, with full image contrasts are presented. More detailed evaluation results are shown in Extended Table E2. The fine-tuned PLM Chinese RoBERTa exhibited the best performance on three out of four tasks, achieving the best average ACC of 0.795 and F1-score of 0.795 across four tasks. The best LLM was the fine-tuned Baichuan2-13B, achieving an average ACC of 0.769 and F1-score of 0.765. Note that the average performance of each language model across the four tasks outperformed or was at least close to the best image-based model Swin Transformer in terms of AUC, ACC, and F1-score. This shows that not only Chinese RoBERTa but in general language models tended to make more reliable brain disease classification than image-based models.

## Discussion

The rapid development of language models has introduced new opportunities for completing practical clinical tasks through the use of medical text information, such as clinical notes^14^. However, there is a lack of studies on brain disease diagnosis via language model analysis of radiological reports, where existing approaches have been based on the brain imaging information^36^. In this work, we have explored advanced language models for classifying brain diseases based on radiological reports, aiming to provide a new paradigm for automated brain disease diagnosis in real clinical scenarios. The results indicate that fine-tuned language models outperform conventional image-based models in terms of classification reliability. Our work has offered a new and effective solution to accurate classification of brain diseases, and it has contributed to the exploration of advanced language models for clinical applications.

The better performance of language models than image-based models can be attributed to the characteristics of the data. First, in terms of data size and dimensionality, natural language data is remarkably smaller than 3D brain image data. The reduction in data complexity makes it easier for the language model to learn the association between reports and diseases. Second, in terms of data content, the radiological reports summarize important information derived from the image, such as descriptions about the anatomical structures and anomalies. Such semantic information alleviates the difficulty in image data understanding.

Different types of popular language models and how to train these models have been investigated in our work. The results shown in Extended Table E2 indicate that the models developed specifically for Chinese language processing, i.e., Chinese RoBERTa and Baichuan2-13B, are better than general multilingual models, RoBERTa-base and LLaMA3-8B, for processing Chinese radiological reports. This observation is consistent with existing studies^25,27^, where models trained on Chinese datasets acquire a better understanding of Chinese. In addition, we have further considered different numbers of parameters and two distinct fine-tuning methods, quantized low-rank adaptation (QLoRA)^37^ and LoRA for the Baichuan2 model. The results presented in Extended Table E4 advocate the joint use of larger parameter size of 13 billion and LoRA fine-tuning.

Among the two kinds of language models, the PLM Chinese RoBERTa performed better than the LLMs (see Extended Table E2). However, the LLMs also have their potential benefits. First, it can provide more efficient solutions to disease classification, as a single pre-trained LLM can perform several different tasks simultaneously, where different PLMs are needed for different tasks. Beyond this, the LLMs can potentially benefit from even larger data volume by simultaneous training with other similar tasks. This can be observed in Extended Table E3, as the average LLM performance is noticeably improved with simultaneous fine-tuning based on all tasks. The LLM performance may be worth further exploration in future work when more tasks are take into account. Furthermore, the LLMs may better adapt to the various writing styles of radiological reports from different hospitals than PLMs, as shown in Extended Table E5, where the LLMs have better performance than the PLMs for the two external hospitals in terms of ACC and F1-score. This may be attributed to their extensive pre-training on a massive amount of natural language data, which allows better handling of cross-site text variations.

In clinical practice, missing image contrasts are pretty common for patients due to their various conditions, and the problem has long been a serious challenge for real-world applications of image-based disease classification approaches^1^. In addition, it is also challenging to generalize an image-based classification model to an unseen site^38^, where the performance can degrade drastically due to domain shift^39^. Our results reveal that the language models are better than image-based models at addressing missing image contrasts and cross-cite data variability. The better ability of the language models at addressing missing image contrasts and cross-site variations may be attributed to the following reasons. First, the language models are not constrained by image contrasts and can focus on all available content in the reports for better performance, whereas the synthesised image contrasts may introduce biases for the image-based models. In addition, the cross-site changes in image and report data are different. For the radiological reports, the changes are generally in writing style, while the anatomical structures and anomalies of greater significance remain consistent. However, the changes in imaging data, including image quality, signal intensity, and others, can noticeably influence the effectiveness of image-based models.

Our study has limitations. First, the sample size in the experiments could still be enlarged. As it may be costly to acquire the gold standard diagnosis results, each task included thousands of patients. Future work will further accumulate the data and increase the sample size by orders of magnitude for a more extensive evaluation. Such increase in the sample size may also improve the performance of language models. Second, the comparison between PLMs and LLMs may be limited by the sample size we have collected. LLMs tend to be more effective given a huge amount of data, and it is still unknown whether LLMs will outperform PLMs given substantially more training data. Finally, in the current model implementation, there is a lack of explainability about the model’s decision. It would be interesting to explore explainable artificial intelligence techniques for the language models to provide better confidence about the diagnosis results.

In conclusion, our study highlights the potential of language models for accurate brain disease diagnosis based on radiological reports. The language model achieved better diagnostic performance than conventional image-based models. Furthermore, the language model is robust to imperfect data conditions, such as missing image contrasts and cross-site variations. Looking forward, our research can contribute to the improvement of diagnostic techniques for brain diseases and the exploration of application of language models in the medical context.

## Methods

### Datasets preparation and pre-processing

Four different types of brain disease classification tasks were considered. We initially collected a total number of 17,507 patients and then filtered the patients based on the presence of image contrasts and quality of radiological reports. The detailed filtering procedure for each task is shown in Figure 3. For each task, patients with poor report quality or those with preoperative preparation reports were first excluded. Then, patients were selected according to their absence of image contrasts. In this work, patients with all five image contrasts were first selected, from which the training set were derived. For the remaining patients, only those with the most common types of missing image contrasts were considered, while those with any other types of missing image contrasts were excluded. For the task of IDH genotyping, patients that did not have FLAIR images only, ADC images only, or both FLAIR and ADC images from Beijing Tiantan Hospital were included. For the two external hospitals, patients from The First Affiliated Hospital of Xinjiang Medical University who had all five image contrasts were included, while patients from Huashan Hospital, Fudan University who did not have T2w images only were included. For the other three tasks, only patients with missing FLAIR images were kept. Our final datasets comprised 14,970 patients, each associated with MRI scans and the corresponding radiological reports.

**Figure 3.**
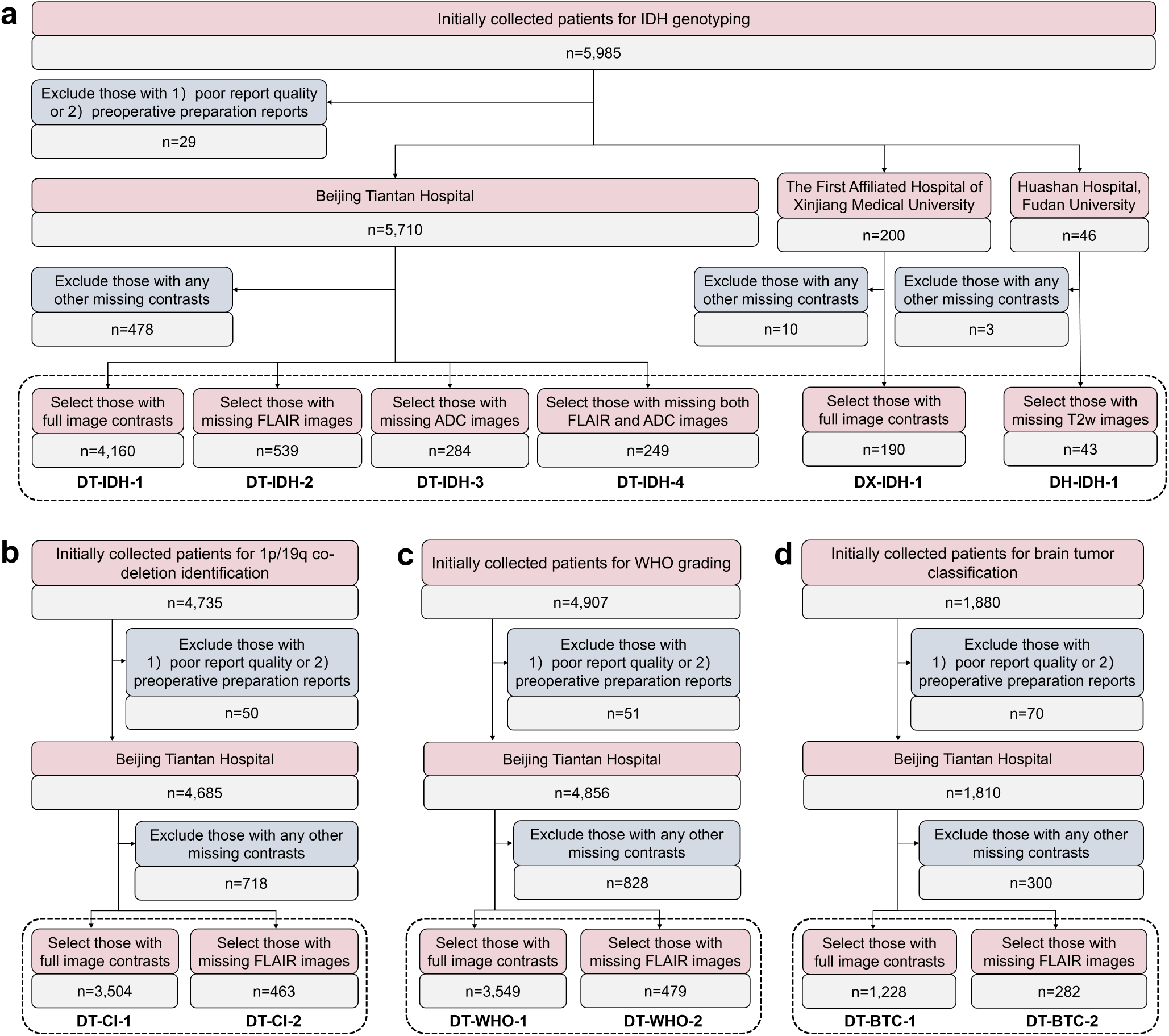
The flowchart of data filtering for the four brain disease classification tasks: (a) IDH genotyping, (b) 1p/19q co-deletion identification, (c) WHO grading, and (d) brain tumor classification. The final included patients for each task are indicated by black dotted boxes.

For image data pre-processing, N4 bias field correction was first applied with a shrink factor of 2 with the Advanced Normalization Tools (ANTs) software^40^. Subsequently, all image contrasts were co-registered to the T1w image. The T1w image was affinely registered to the MNI152 template^41^ with linear interpolation. The transformation matrix was then applied to the other image contrasts so that they were all aligned with the MNI152 template. Finally, the brain mask was extracted for skull striping with ROBEX^42^. Each pre-processed image had a size of 256×256×256 with a resolution of 1 mm^3^. For report data pre-processing, nonsensical characters were first removed from each report. Then, the reports were tokenized and either padded or truncated to a maximum length of 256 to as model input.

For image-based brain disease classification, additional tumor segmentation models and image synthesis models were obtained. The tumor segmentation was achieved with a nnU-Net model^43^, which was trained on 1500 patients from the BraTS2021 dataset^44^ for whole tumor segmentation. The missing image contrast synthesis was accomplished with a pGAN model^35^, which was individually trained for each missing contrast and each task with 100 patients with full image contrasts randomly selected from the training set of DT-IDH-1, DT-CI-1, DT-WHO-1, and DT-BTC-1. All models used in our experiments were trained with two NVIDIA RTX A6000 GPUs.

### The PLMs for brain disease classification

PLMs, such as BERT-based models, have achieved excellent performance on multiple natural language processing tasks by pre-training on a diverse corpus of language understanding tasks and subsequently fine-tuning on specific tasks. Derivatives of BERT, such as RoBERTa^24^, have further advanced domain-specific natural language processing technologies. In this work, the general RoBERTa-base model^24^ with a 12-layer architecture and approximately 125 million parameters was adopted. Further, considering that our collected radiological reports were written in Chinese, the Chinese RoBERTa model, which employs whole word masking for Chinese word segmentation, has been taken into account. Specifically, we adopted the Chinese RoBERTa-wwm^25^ pre-trained on additional EXT data^25^.

We fine-tuned the models in a supervised manner using the training and validation sets of DT-IDH-1, DT-CI-1, DT-WHO-1, and DT-BTC-1. The fine-tuning was performed for each task independently based on the radiological report and classification label of each patient, where the cross-entropy loss function^45^ was minimized. The Adam optimizer^46^ was used with a batch size of eight and 20 epochs for training convergence. Model selection was performed based on the smallest cross-entropy loss of the validation set. Taking the class imbalance of the training data into consideration, class weights were applied to the standard loss function to assign higher weights to minority classes and lower weights to majority classes.

### The LLMs for brain disease classification

The generative LLMs, such as GPT^47^, T5^48^, and LLaMA^49^, have acquired vast knowledge across multiple specialized domains through pre-training and have demonstrated efficient transfer learning capabilities for new tasks. In this work, we employed the generative LLMs to predict the classification of brain diseases. Due to constraints imposed by medical data privacy, only openly accessible base models were selected for our local deployment. Specifically, the latest version of LLaMA (version 3)^26^ with 8 billion parameters (referred to as LLaMA3-8B) was adopted, as it had been shown to outperform many openly accessible LLMs on common industry benchmarks. In addition, Baichuan (version 2)^27^ with 13 billion parameters (referred to as Baichuan2-13B) was adopted, as it had achieved the best performance on multiple benchmarks in both Chinese and English. Since the LLMs do not have category limitations on their outputs, we fine-tuned the LLMs using training data from all four tasks simultaneously. The training data combined the training sets of DT-IDH-1, DT-CI-1, DT-WHO-1, and DT-BTC-1, and different tasks were distinguished by different prompts. The detailed prompt for each task is shown in Table 4. Each prompt included three parts, the radiological report of each patient, role assignment as a radiologist, and a question asking the model to generate a single precise classification label. The parameter efficient fine-tuning method low-rank adaptation (LoRA)^28^ was used with a batch size of 4, gradient accumulation steps of 4, a rank of 16, an alpha value of 32, and a temperature value of 1.

**Table 4.**
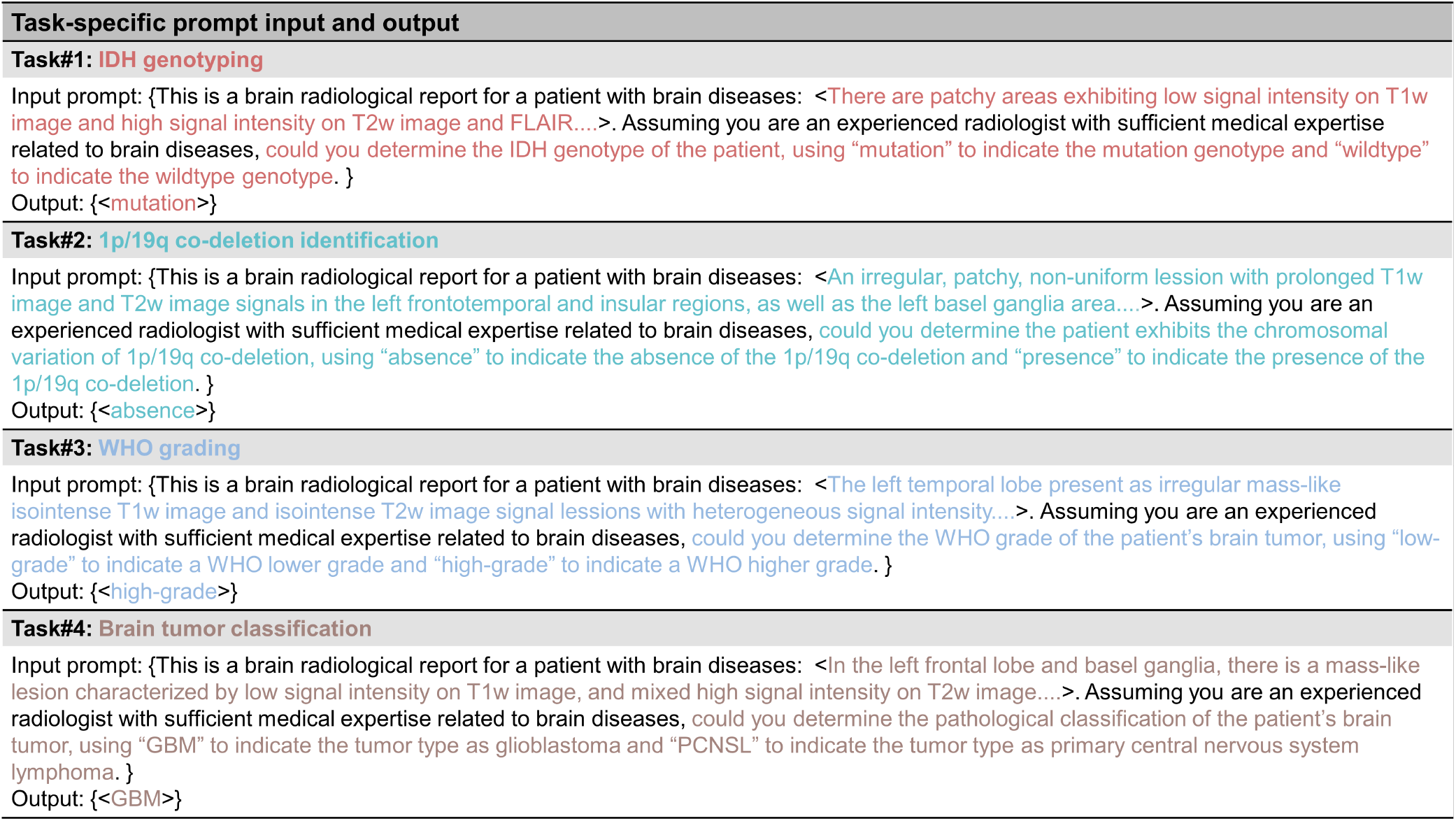
Examples of the input prompt and output setting for fine-tuning the LLMs for each task. The contents within the brackets <> represent the radiological reports of different patients and their corresponding disease labels, while the other colored contents indicate task-specific prompt settings.

Each model was trained for 3 epochs with an initial learning rate of 5e-5, which was adjusted according to a cosine annealing schedule.

### Competing image-based models

We considered six conventional image-based classification models for comparison, including those specifically designed for brain disease classification and the latest widely used models in the general domain. The models included those that process imaging data in 2D, 2.5D, and 3D formats, and some of these classification models were aided by the tumor segmentation results. Their detailed description is given below

- **DeepRisk**. The DeepRisk^29^ model is developed based on a 2D ResNet34^50^ backbone and several attention^51^ blocks. This model was designed to make predictions directly using whole-brain MRI scans without tumor segmentation. Specifically, we took eight equidistant slices from each MRI contrast and concatenated them for classification.
- **2D & 3D MedMNIST**. The MedMNIST^30^ provides benchmarks for 2D and 3D biomedical image classification based on a 2D ResNet^50^ and a 3D ResNet^52^, respectively. For the 2D-based architecture **2D MedMNIST**, we took the slice with the largest tumor area from each MRI sequence and concatenated them for classification. For the 3D-based architecture **3D MedMNIST**, the whole-brain MRI scans were directly used for classification.
- **DenseNet**. The DenseNet^31^ model, designed based on stacked dense blocks^53^, performed disease classification based on the images and tumor segmentation. In addition to the slice with the largest tumor area, its input concatenated eleven slices before and twelve slices after this slice for classification, along with the slice itself.
- **ViT**. The pre-trained Vision Transformer (ViT)^32^ has learned image features from extensive imaging data, achieving breakthrough performance on a variety of vision-related tasks in the general domain. We adopted the ViT model of base size that was pre-trained on ImageNet-21k^54^ at a resolution of 224×224. The model input comprised the slice with the largest tumor area from each MRI contrast and subsequently concatenated them for classification.
- **Swin Transformer**. Swin Transformer^33^ is a recently proposed Transformer-based vision model that uses shifted windows to capture local features in images, while also ensuring computational efficiency. We adopted Swin Transformer V2 of tiny size pre-trained on ImageNet-1k^54^ at a resolution of 256×256. The model input was identical to that of ViT in our experiments.

## Supporting information

Supplementary Information of Simple Words over Rich Imaging: Accurate Brain Disease Classification via Language Model Analysis of Radiological Reports

## Data Availability

All data produced in the present study are available upon reasonable request to the authors.
All data produced in the present work are contained in the manuscript.

## Acknowledgements

This study was supported by the Beijing Municipal Natural Science Foundation (7242273 & JQ20035), Fundamental Research Funds for the Central Universities (2022CX11008), Xiaomi Young Scholars Program, National Natural Science Foundation of China (81870958 & 81571631), and Special Fund of the Pediatric Medical Coordinated Development Center of Beijing Hospitals Authority (XTYB201831).

## Author contributions statement

Xin Gao wrote the code, conducted experiments, and wrote the manuscript. Jun Qiu, Tiantian Hua and Ying Jin interpreted the data and made diagnoses. Xin Gao, Longfei Chen, Shanbo Zhao, Zhiqiang Wu and Haotian Hou participated in model design and optimization. Junjie Li, Yunling Wang, Wei Zhao and Yuxin Li collected the data and provided access to it. Jun Qiu, Junjie Li, Xuzhu Chen, Yunyun Duan and Yaou Liu offered guidance in medical expertise. Meihui Zhang and Chuyang Ye conceived, designed and directed the study. All authors reviewed, edited and approved the manuscript.

## Supplementary materials

Supplementary information is available for this paper.

**Table E1.** The pre-processing time costs for image and report data.

| Data | Pre-processing item | Average time cost (s) | Total cost (s) |
| --- | --- | --- | --- |
| Image | N4-bias field correction | 52.678 | 159.978 |
|  | Image co-registration | 58.160 |  |
|  | Skull stripping | 23.379 |  |
|  | Tumor segmentation | 25.761 |  |
| Report | Removal nonsensical characters,<br>tokenization,<br>padding,<br>truncating. | 0.026 |  |

**Table E2.** The detailed comparison results of four language models across four tasks for patients with full image contrasts from datasets DT-IDH-1, DT-CI-1, DT-WHO-1, and DT-BTC-1. The best results are highlighted in bold.

| Model type | Model | AUC | ACC | F1-score | SEN | SPEC | PPV | NPV |
| --- | --- | --- | --- | --- | --- | --- | --- | --- |
| IDH genotyping (DT-IDH-1) |  |  |  |  |  |  |  |  |
| PLMs | RoBERTa-base | 0.819($\pm 0.010$) | 0.753($\pm 0.005$) | 0.754($\pm 0.006$) | 0.754($\pm 0.010$) | 0.753($\pm 0.017$) | 0.788($\pm 0.010$) | 0.716($\pm 0.006$) |
| | Chinese RoBERTa | **0.863($\pm 0.005$)** | **0.807($\pm 0.009$)** | **0.807($\pm 0.010$)** | **0.830($\pm 0.017$)** | 0.779($\pm 0.036$) | **0.821($\pm 0.021$)** | 0.790($\pm 0.010$) |
| LLMs | LLaMA3-8B | - | 0.782 | 0.782 | 0.750 | **0.808** | 0.762 | **0.797** |
|  | Baichuan2-13B | - | 0.770 | 0.770 | 0.755 | 0.781 | 0.739 | 0.795 |
| 1p/19q co-deletion identification (DT-CI-1) |  |  |  |  |  |  |  |  |
| PLMs | RoBERTa-base | 0.739($\pm 0.012$) | 0.682($\pm 0.014$) | 0.693($\pm 0.012$) | 0.642($\pm 0.038$) | 0.700($\pm 0.030$) | 0.474($\pm 0.017$) | 0.824($\pm 0.011$) |
| | Chinese RoBERTa | **0.763($\pm 0.007$)** | **0.747($\pm 0.012$)** | **0.749($\pm 0.010$)** | 0.600($\pm 0.015$) | **0.809($\pm 0.017$)** | 0.570($\pm 0.021$) | **0.828($\pm 0.005$)** |
| LLMs | LLaMA3-8B | - | 0.676 | 0.666 | **0.806** | 0.365 | **0.752** | 0.442 |
|  | Baichuan2-13B | - | 0.645 | 0.631 | 0.794 | 0.288 | 0.727 | 0.370 |
| WHO grading (DT-WHO-1) |  |  |  |  |  |  |  |  |
| PLMs | RoBERTa-base | 0.846($\pm 0.009$) | 0.773($\pm 0.008$) | 0.770($\pm 0.007$) | **0.845($\pm 0.013$)** | 0.634($\pm 0.009$) | 0.802($\pm 0.003$) | 0.715($\pm 0.018$) |
| | Chinese RoBERTa | **0.874($\pm 0.002$)** | **0.804($\pm 0.009$)** | **0.804($\pm 0.008$)** | 0.842($\pm 0.025$) | **0.737($\pm 0.020$)** | **0.847($\pm 0.007$)** | **0.731($\pm 0.025$)** |
| LLMs | LLaMA3-8B | - | 0.771 | 0.772 | 0.811 | 0.703 | 0.826 | 0.682 |
|  | Baichuan2-13B | - | 0.789 | 0.789 | 0.833 | 0.711 | 0.833 | 0.711 |
| Brain tumor classification (DT-BTC-1) |  |  |  |  |  |  |  |  |
| PLMs | RoBERTa-base | 0.822($\pm 0.015$) | 0.765($\pm 0.014$) | 0.747($\pm 0.021$) | 0.445($\pm 0.073$) | **0.911($\pm 0.013$)** | 0.694($\pm 0.007$) | 0.784($\pm 0.019$) |
| | Chinese RoBERTa | **0.890($\pm 0.009$)** | 0.821($\pm 0.021$) | 0.818($\pm 0.018$) | 0.665($\pm 0.048$) | 0.893($\pm 0.048$) | 0.752($\pm 0.082$) | 0.854($\pm 0.012$) |
| LLMs | LLaMA3-8B | - | 0.781 | 0.776 | 0.875 | 0.575 | 0.819 | 0.676 |
|  | Baichuan2-13B | - | **0.875** | **0.871** | **0.955** | 0.700 | **0.875** | **0.875** |

**Table E3.** The comparison results of fine-tuning the LLM Baichuan2-13B with all tasks simultaneously or with each task respectively.

| Fine-tuning type | Average performance |  | IDH genotyping |  | 1p/19q co-deletion identification |  | WHO grading |  | Brain tumor classification |  |
| --- | --- | --- | --- | --- | --- | --- | --- | --- | --- | --- |
|  | ACC | F1-score | ACC | F1-score | ACC | F1-score | ACC | F1-score | ACC | F1-score |
| All tasks were fine-tuned simultaneously | 0.770 | 0.765 | 0.770 | 0.770 | 0.645 | 0.631 | 0.789 | 0.789 | 0.875 | 0.871 |
| Each task was fine-tuned respectively | 0.733 | 0.723 | 0.750 | 0.750 | 0.653 | 0.617 | 0.794 | 0.794 | 0.734 | 0.730 |

**Table E4.** The comparison results for the Baichuan2 model with different numbers of parameters and fine-tuning methods. The average performance was calculated from four tasks for each model.

| Base model | Parameter | Fine-tuning type | Average performance |  | IDH genotyping |  | 1p/19q co-deletion identification |  | WHO grading |  | Brain tumor classification |  |
| --- | --- | --- | --- | --- | --- | --- | --- | --- | --- | --- | --- | --- |
|  |  |  | ACC | F1-score | ACC | F1-score | ACC | F1-score | ACC | F1-score | ACC | F1-score |
| Baichuan2 | 7B | QLoRA | 0.734 | 0.714 | 0.757 | 0.756 | 0.670 | 0.607 | 0.749 | 0.736 | 0.758 | 0.755 |
| Baichuan2 | 7B | LoRA | 0.757 | 0.754 | 0.782 | 0.782 | 0.656 | 0.643 | 0.794 | 0.793 | 0.797 | 0.797 |
| Baichuan2 | 13B | QLoRA | 0.760 | 0.759 | 0.752 | 0.752 | 0.636 | 0.639 | 0.783 | 0.779 | 0.867 | 0.867 |
| Baichuan2 | 13B | LoRA | **0.770** | **0.765** | 0.770 | 0.770 | 0.645 | 0.631 | 0.789 | 0.789 | 0.875 | 0.871 |

**Table E5.** The detailed comparison results of two kinds of language models, PLMs and LLMs, using two external datasets, DX-IDH-1 and DH-IDH-1, from The First Affiliated Hospital of Xinjiang Medical University and Huashan Hospital, Fudan University, for the task of IDH genotyping.

| Model type | Model | DX-IDH-1 |  |  | DH-IDH-1 |  |  |
| --- | --- | --- | --- | --- | --- | --- | --- |
|  |  | AUC | ACC | F1-score | AUC | ACC | F1-score |
| PLMs | RoBERTa-base | 0.762(±0.011) | 0.757(±0.014) | 0.728(±0.023) | 0.837(±0.015) | 0.739(±0.017) | 0.727(±0.022) |
|  | Chinese RoBERTa | **0.865(±0.012)** | 0.815(±0.011) | 0.807(±0.012) | **0.848(±0.023)** | 0.772(±0.022) | 0.765(±0.024) |
| LLMs | LLaMA3-8B | - | **0.821** | **0.810** | - | **0.790** | **0.790** |
|  | Baichuan2-13B | - | 0.778 | 0.770 | - | **0.790** | 0.785 |

