## Supplementary Information of Simple Words over Rich Imaging: Accurate Brain Disease Classification via Language Model Analysis of Radiological Reports for "Simple Words over Rich Imaging: Accurate Brain Disease Classification via Language Model Analysis of Radiological Reports"

### Implementation details of our language models and competing image-based models

The language models used the average representation of the sequence for classification, where the hidden size of the final fully connected layer was 768. For language model training, both the RoBERTa-base [6] model and the Chinese RoBERTa [1] model adopted the cross-entropy loss [10] with respective loss weights for each task. The training loss weight for each class was set inversely proportional to its numbers of training samples. Specifically, the loss weights for the four tasks were 1.21/1.00 (IDH wildtype/mutation), 1.00/2.47 (1p/19q codeletion absence/presence), 1.72/1.00 (WHO low-grade/high-grade), and 1.00/2.28 (GBM/PCNSL), respectively.

The implementation of the six image-based models followed the settings described in their original papers. For the DeepRisk [9] model, the cross-entropy loss was adopted, and the hidden size of the final fully connected layer was 512. The input image size for DeepRisk was [40, 256, 256], which was obtained by sampling 8 equidistant slices from each of the five image contrasts. For the 2D MedMNIST [8] model, the hidden size of the final fully connected layer was 512. The input image size was [5, 256, 256], where the slices with the largest tumor area from the five MRI contrasts were concatenated. For the 3D MedMNIST [8] model, the hidden size of the final fully connected layer was 512. The input image size was [5, 256, 256, 256], which included the original five image contrasts. For the DenseNet [4] model, the hidden size of the final fully connected layer was 1,664. The model processed each image contrast separately and computed the average representation of the five image contrasts for classification. The input size of each image contrast was [24, 256, 256], where the 24 slices included eleven slices before and twelve slices after the slice with the largest tumor area, along with the slice itself. For both the ViT [2] the Swin Transformer [7] model, the hidden size of the final fully connected layer was 512. The input image size was [5, 256, 256], where the models took the slice with the largest tumor area from each MRI contrast and concatenated these slices for classification.

### Details of imaging data pre-processing

For the imaging data, the patients were first categorized based on the availability of image contrasts. For all selected patients, the T1c image contrast was included, and brain tumor segmentation was performed based on the T1c image. The N4 bias field correction was applied first. Next, registration was performed between the T1w image and the MIN152 template [3] using affine transformation and linear interpolation. The obtained affine matrix was then applied to other image contrasts. Finally, skull striping was performed based on the T1w image using ROBEX [5].

### Detailed classification results of our language models and competing image-based models

First, for the four tasks with full image contrasts (associated with DT-IDH-1, DT-CI-1, DT-WHO-1, and DT-BTC-1), we provide the specific confusion matrices of the language model Chinese RoBERTa and six image-based models. The results are illustrated in Supplementary Figure S1. The detailed results demonstrate the superior classification performance of Chinese RoBERTa compared to the image-based models, where the language model exhibited minimal bias across different categories and achieved the highest correct classification counts in nearly all categories.

Then, for the experiments with missing image contrasts (associated with DT-IDH-2, DT-IDH-3, DT-IDH-4, DT-CI-2, DT-WHO-2, and DT-BTC-2), we first supplement the classification results of the language model and image-based models with additional metrics that are not available in the main text. The results are presented in Supplementary Table S1, where the AUC, ACC, F1-score, SEN, SPEC, PPV, and NPV are reported. The detailed results indicate that Chinese RoBERTa generally outperformed the competing image-based models and better addressed the issue of missing image contrasts. Moreover, we present the detailed classification results of all four language models for the experiments with missing image contrasts in Supplementary Table S2. The Chinese RoBERTa performed best overall, achieving the highest performance in five of the six datasets.

In addition, we present the detailed classification results for the two external datasets DX-IDH-1 and DH-IDH-1. The results are shown in Supplementary Table S3, where the AUC, ACC, F1-score, SEN, SPEC, PPV, and NPV are reported. The results support the observation that the advantages of the language model over the image-based models were reliable and that it better handled cross-site data variability.

### The detailed comparison with the radiologists

We present the numerical classification results of the language model Chinese RoBERTa, the image-based models, and the evaluation given by the three radiologists. The comparison is shown in Supplementary Table S4, where the AUC, ACC, and F1-score are reported. The comparison between the radiologists, image-based models, and report-based model in terms of the F1-score is consistent with the performance in terms of the ACC, showing that our report-based model outperformed junior radiologists and image-based models, while the experienced radiologist (with ten years of experience) achieved the best performance.

### Data examples of patients from different datasets

To give a more straightforward understanding of the radiological reports of patients with full image contrasts, with missing image contrasts, and from different hospitals more clearly, we provide representative samples from all datasets in Supplementary Table S5. The difference in the number of image contrasts leads to variations in the report length and information richness. Yet the language model effectively addressed these variations caused by missing image contrasts.

In addition, the writing styles of different hospitals were apparently different, particularly in the descriptions of the names of various image contrasts (which were originally in Chinese), as well as in whether the report included descriptions of normal structures and the order of descriptions. Our language model was capable of adapting to the cross-site data variability, as indicated by the results in the main text and supplementary materials.

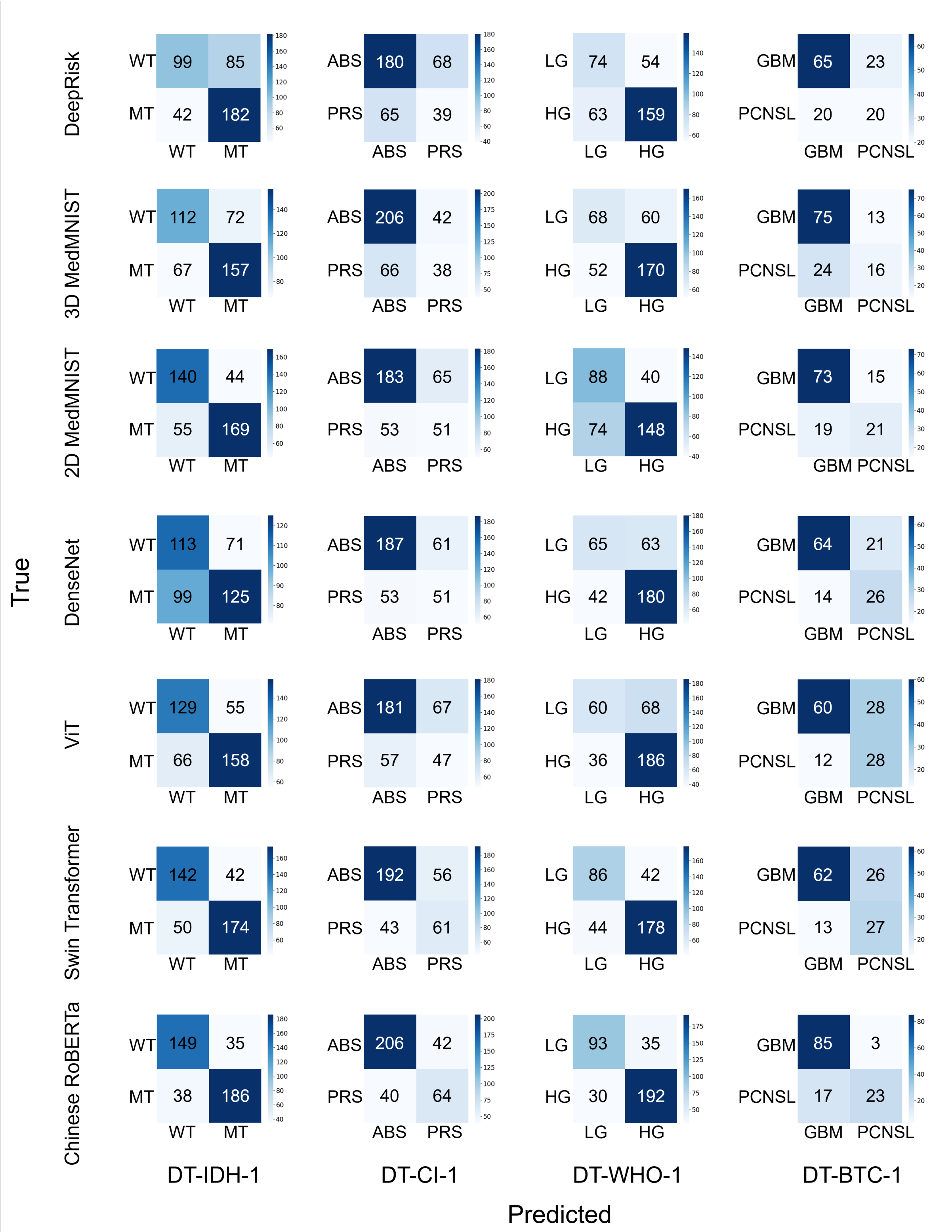


Figure S1: The confusion matrices of Chinese RoBERTa and the image-based models. The vertical axis represents the true labels and the horizontal axis represents the predicted results. All four tasks with full image contrasts on DT-IDH-1, DT-CI-1, DT-WHO-1, and DT-BTC-1 are included. WT: wildtype; MT: mutation; ABS: absence; PRS: presence; LG: low-grade; HG: high-grade.

Table S1: Detailed classification performance of Chinese RoBERTa and the image-based models on DT-IDH-2, DT-IDH-3, DT-IDH-4, DT-CI-2, DT-WHO-2, and DT-BTC-2 for patients with missing image contrasts. The best result is highlighted in bold.

| Data type | Model type | Model | DT-IDH-2 | | | | | | |
|---|---|---|---|---|---|---|---|---|---|
| | | | AUC | ACC | F1-score | SEN | SPEC | PPV | NPV |
| Image | w/o segmentation | DeepRisk | 0.616(±0.011) | 0.600(±0.015) | 0.593(±0.018) | 0.698(±0.047) | 0.485(±0.068) | 0.612(±0.018) | 0.583(±0.018) |
| | | 3D MedMNIST | 0.586(±0.033) | 0.559(±0.020) | 0.553(±0.024) | 0.658(±0.036) | 0.444(±0.045) | 0.578(±0.017) | 0.529(±0.026) |
| | w/ segmentation | 2D MedMNIST | 0.710(±0.032) | 0.656(±0.026) | 0.654(±0.026) | 0.631(±0.055) | 0.684(±0.090) | 0.704(±0.046) | 0.616(±0.025) |
| | | DenseNet | 0.572(±0.012) | 0.528(±0.017) | 0.522(±0.022) | 0.438(±0.077) | 0.632(±0.052) | 0.578(±0.009) | 0.495(±0.015) |
| | | ViT | 0.722(±0.013) | 0.664(±0.013) | 0.663(±0.013) | 0.663(±0.053) | 0.665(±0.058) | 0.698(±0.022) | 0.633(±0.021) |
| | | Swin Transformer | 0.769(±0.006) | 0.707(±0.009) | 0.707(±0.001) | 0.712(±0.004) | 0.701(±0.022) | 0.734(±0.014) | 0.678(±0.006) |
| Radiological report | our model | Chinese RoBERTa | **0.808(±0.005)** | **0.753(±0.010)** | **0.753(±0.010)** | **0.759(±0.032)** | **0.745(±0.017)** | **0.775(±0.005)** | **0.729(±0.023)** |
| **Data type** | **Model type** | **Model** | **DT-IDH-3** | | | | | | |
| | | | **AUC** | **ACC** | **F1-score** | **SEN** | **SPEC** | **PPV** | **NPV** |
| Image | w/o segmentation | DeepRisk | 0.568(±0.021) | 0.680(±0.022) | 0.614(±0.017) | **0.911(±0.036)** | 0.090(±0.039) | 0.718(±0.008) | 0.300(±0.082) |
| | | 3D MedMNIST | 0.502(±0.015) | 0.623(±0.054) | 0.601(±0.022) | 0.777(±0.118) | 0.230(±0.111) | 0.719(±0.005) | 0.299(±0.031) |
| | w/ segmentation | 2D MedMNIST | 0.615(±0.030) | 0.659(±0.015) | 0.658(±0.013) | 0.762(±0.036) | 0.395(±0.064) | 0.763(±0.012) | 0.394(±0.025) |
| | | DenseNet | 0.598(±0.007) | 0.609(±0.029) | 0.620(±0.021) | 0.676(±0.065) | 0.440(±0.068) | 0.755(±0.009) | 0.349(±0.017) |
| | | ViT | 0.648(±0.018) | 0.649(±0.033) | 0.660(±0.028) | 0.700(±0.055) | 0.520(±0.051) | 0.788(±0.014) | 0.407(±0.035) |
| | | Swin Transformer | 0.632(±0.029) | 0.645(±0.029) | 0.655(±0.026) | 0.702(±0.049) | 0.500(±0.070) | 0.782(±0.020) | 0.398(±0.036) |
| Radiological report | our model | Chinese RoBERTa | **0.780(±0.009)** | **0.734(±0.010)** | **0.742(±0.007)** | 0.767(±0.025) | **0.650(±0.030)** | **0.848(±0.007)** | **0.524(±0.016)** |
| **Data type** | **Model type** | **Model** | **DT-IDH-4** | | | | | | |
| | | | **AUC** | **ACC** | **F1-score** | **SEN** | **SPEC** | **PPV** | **NPV** |
| Image | w/o segmentation | DeepRisk | 0.516(±0.039) | 0.542(±0.017) | 0.462(±0.011) | **0.892(±0.052)** | 0.135(±0.030) | 0.545(±0.008) | 0.547(±0.100) |
| | | 3D MedMNIST | 0.489(±0.022) | 0.522(±0.024) | 0.503(±0.014) | 0.694(±0.086) | 0.323(±0.057) | 0.543(±0.014) | 0.483(±0.038) |
| | w/ segmentation | 2D MedMNIST | 0.544(±0.025) | 0.539(±0.027) | 0.527(±0.034) | 0.673(±0.046) | 0.384(±0.086) | 0.561(±0.025) | 0.498(±0.039) |
| | | DenseNet | 0.619(±0.012) | 0.573(±0.020) | 0.566(±0.020) | 0.619(±0.122) | 0.520(±0.119) | 0.603(±0.017) | 0.546(±0.032) |
| | | ViT | 0.606(±0.004) | 0.593(±0.022) | 0.591(±0.021) | 0.637(±0.061) | 0.542(±0.057) | 0.619(±0.021) | 0.564(±0.027) |
| | | Swin Transformer | 0.596(±0.017) | 0.594(±0.015) | 0.593(±0.015) | 0.629(±0.025) | 0.553(±0.037) | 0.622(±0.017) | 0.561(±0.016) |
| Radiological report | our model | Chinese RoBERTa | **0.722(±0.008)** | **0.682(±0.009)** | **0.681(±0.009)** | 0.743(±0.013) | **0.612(±0.027)** | **0.691(±0.012)** | **0.671(±0.008)** |
| **Data type** | **Model type** | **Model** | **DT-CI-2** | | | | | | |
| | | | **AUC** | **ACC** | **F1-score** | **SEN** | **SPEC** | **PPV** | **NPV** |
| Image | w/o segmentation | DeepRisk | 0.625(±0.011) | 0.665(±0.029) | 0.676(±0.019) | 0.469(±0.073) | 0.729(±0.062) | 0.368(±0.023) | 0.807(±0.009) |
| | | 3D MedMNIST | 0.578(±0.035) | 0.547(±0.032) | 0.576(±0.029) | 0.544(±0.069) | 0.548(±0.050) | 0.284(±0.023) | 0.784(±0.022) |
| | w/ segmentation | 2D MedMNIST | 0.603(±0.011) | 0.676(±0.007) | 0.678(±0.010) | 0.370(±0.074) | **0.778(±0.026)** | 0.352(±0.027) | 0.789(±0.014) |
| | | DenseNet | 0.652(±0.010) | 0.684(±0.028) | 0.690(±0.018) | 0.455(±0.083) | 0.759(±0.065) | 0.390(±0.023) | 0.809(±0.011) |
| | | ViT | 0.563(±0.010) | 0.605(±0.026) | 0.625(±0.021) | 0.448(±0.044) | 0.656(±0.046) | 0.302(±0.016) | 0.782(±0.007) |
| | | Swin Transformer | 0.670(±0.009) | 0.666(±0.013) | 0.682(±0.011) | 0.532(±0.021) | 0.710(±0.015) | 0.378(±0.016) | 0.821(±0.007) |
| Radiological report | our model | Chinese RoBERTa | **0.728(±0.004)** | **0.705(±0.007)** | **0.716(±0.006)** | **0.546(±0.031)** | 0.758(±0.015) | **0.427(±0.010)** | **0.834(±0.007)** |
| **Data type** | **Model type** | **Model** | **DT-WHO-2** | | | | | | |
| | | | **AUC** | **ACC** | **F1-score** | **SEN** | **SPEC** | **PPV** | **NPV** |
| Image | w/o segmentation | DeepRisk | 0.655(±0.011) | 0.605(±0.024) | 0.618(±0.023) | 0.577(±0.040) | 0.664(±0.047) | 0.786(±0.020) | 0.425(±0.022) |
| | | 3D MedMNIST | 0.576(±0.021) | 0.553(±0.031) | 0.566(±0.031) | 0.526(±0.068) | 0.610(±0.050) | 0.742(±0.006) | 0.378(±0.017) |
| | w/ segmentation | 2D MedMNIST | 0.626(±0.016) | 0.613(±0.031) | 0.622(±0.027) | 0.646(±0.056) | 0.542(±0.053) | 0.751(±0.018) | 0.421(±0.032) |
| | | DenseNet | 0.737(±0.006) | 0.706(±0.012) | 0.702(±0.009) | 0.795(±0.056) | 0.516(±0.091) | 0.780(±0.023) | 0.546(±0.024) |
| | | ViT | 0.751(±0.015) | 0.685(±0.018) | 0.693(±0.016) | 0.677(±0.055) | **0.702(±0.095)** | 0.832(±0.033) | 0.507(±0.020) |
| | | Swin Transformer | 0.772(±0.005) | 0.709(±0.013) | 0.706(±0.014) | 0.800(±0.015) | 0.051(±0.045) | 0.779(±0.014) | 0.547(±0.021) |
| Radiological report | our model | Chinese RoBERTa | **0.861(±0.003)** | **0.800(±0.004)** | **0.797(±0.004)** | **0.876(±0.011)** | 0.639(±0.013) | **0.838(±0.003)** | **0.708(±0.015)** |
| **Data type** | **Model type** | **Model** | **DT-BTC-2** | | | | | | |
| | | | **AUC** | **ACC** | **F1-score** | **SEN** | **SPEC** | **PPV** | **NPV** |
| Image | w/o segmentation | DeepRisk | 0.510(±0.043) | 0.594(±0.071) | 0.593(±0.059) | 0.402(±0.044) | 0.688(±0.124) | 0.409(±0.080) | 0.697(±0.027) |
| | | 3D MedMNIST | 0.620(±0.009) | 0.524(±0.059) | 0.532(±0.056) | 0.612(±0.076) | 0.481(±0.125) | 0.375(±0.041) | 0.714(±0.014) |
| | w/ segmentation | 2D MedMNIST | 0.711(±0.038) | 0.693(±0.044) | 0.689(±0.034) | 0.512(±0.098) | 0.779(±0.107) | 0.559(±0.080) | 0.768(±0.016) |
| | | DenseNet | 0.685(±0.021) | 0.675(±0.015) | 0.667(±0.019) | 0.443(±0.092) | 0.789(±0.044) | 0.508(±0.025) | 0.743(±0.023) |
| | | ViT | 0.636(±0.037) | 0.605(±0.047) | 0.611(±0.041) | 0.509(±0.053) | 0.651(±0.080) | 0.425(±0.052) | 0.729(±0.023) |
| | | Swin Transformer | 0.722(±0.075) | 0.671(±0.055) | 0.678(±0.053) | 0.612(±0.080) | 0.700(±0.054) | 0.503(±0.069) | 0.786(±0.043) |
| Radiological report | our model | Chinese RoBERTa | **0.855(±0.012)** | **0.796(±0.016)** | **0.792(±0.021)** | **0.640(±0.091)** | **0.873(±0.024)** | **0.714(±0.015)** | **0.833(±0.030)** |

Table S2: Detailed comparison of the four language models for patients with missing image contrasts based on DT-IDH-2, DT-IDH-3, DT-IDH-4, DT-CI-2, DT-WHO-2, and DT-BTC-2. The best result is highlighted in bold.

| Model Type | Model | AUC | ACC | F1-score |
|---|---|---|---|---|
| IDH genotyping (DT-IDH-2) | | | | |
| PLMs | RoBERTa-base | 0.770(±0.007) | 0.702(±0.016) | 0.701(±0.016) |
| | Chinese RoBERTa | **0.808(±0.005)** | **0.753(±0.010)** | **0.753(±0.010)** |
| LLMs | LLaMA3-8B | - | 0.717 | 0.716 |
| | Baichuan2-13B | - | 0.706 | 0.706 |
| IDH genotyping (DT-IDH-3) | | | | |
| PLMs | RoBERTa-base | 0.741(±0.012) | 0.718(±0.016) | 0.726(±0.013) |
| | Chinese RoBERTa | **0.780(±0.009)** | **0.734(±0.010)** | **0.742(±0.008)** |
| LLMs | LLaMA3-8B | - | 0.704 | 0.711 |
| | Baichuan2-13B | - | 0.697 | 0.703 |
| IDH genotyping (DT-IDH-4) | | | | |
| PLMs | RoBERTa-base | 0.669(±0.015) | 0.620(±0.009) | 0.620(±0.009) |
| | Chinese RoBERTa | **0.722(±0.008)** | **0.682(±0.009)** | **0.681(±0.009)** |
| LLMs | LLaMA3-8B | - | 0.643 | 0.638 |
| | Baichuan2-13B | - | 0.651 | 0.646 |
| 1p/19q co-deletion identification (DT-CI-2) | | | | |
| PLMs | RoBERTa-base | 0.667(±0.006) | 0.650(±0.010) | 0.668(±0.008) |
| | Chinese RoBERTa | **0.728(±0.004)** | **0.705(±0.007)** | **0.716(±0.006)** |
| LLMs | LLaMA3-8B | - | 0.678 | 0.669 |
| | Baichuan2-13B | - | 0.673 | 0.672 |
| WHO grading (DT-WHO-2) | | | | |
| PLMs | RoBERTa-base | 0.846(±0.009) | 0.773(±0.008) | 0.770(±0.007) |
| | Chinese RoBERTa | **0.861(±0.003)** | **0.800(±0.004)** | **0.797(±0.004)** |
| LLMs | LLaMA3-8B | - | 0.774 | 0.773 |
| | Baichuan2-13B | - | 0.789 | 0.788 |
| Brain tumor classification (DT-BTC-2) | | | | |
| PLMs | RoBERTa-base | 0.822(±0.015) | 0.765(±0.014) | 0.747(±0.021) |
| | Chinese RoBERTa | **0.855(±0.012)** | 0.796(±0.016) | 0.792(±0.021) |
| LLMs | LLaMA3-8B | - | 0.808 | 0.800 |
| | Baichuan2-13B | - | **0.829** | **0.828** |

Table S3: Detailed classification performance of Chinese RoBERTa and the image-based models for the external datasets DX-IDH-1 and DH-IDH-1. The best result is highlighted in bold.

| Data type | Model type | Model | The First Affiliated Hospital of Xinjiang Medical University (DX-IDH-1) | | | | | | | | |
|---|---|---|---|---|---|---|---|---|---|---|---|
| | | | AUC | *p* (AUC) | ACC | *p* (ACC) | F1-score | SEN | SPEC | PPV | NPV |
| Image | w/o segmentation | DeepRisk | 0.612(±0.044) | 7.169E-4 | 0.629(±0.067) | 7.889E-3 | 0.635(±0.057) | 0.869(±0.052) | 0.261(±0.022) | 0.528(±0.018) | 0.687(±0.104) |
| | | 3D MedMNIST | 0.587(±0.043) | 2.406E-4 | 0.603(±0.060) | 1.465E-3 | 0.615(±0.045) | 0.726(±0.020) | 0.500(±0.055) | 0.582(±0.026) | 0.655(±0.032) |
| | w/ segmentation | 2D MedMNIST | 0.597(±0.046) | 4.648E-4 | 0.651(±0.035) | 9.615E-4 | 0.647(±0.022) | 0.228(±0.046) | **0.854(±0.034)** | 0.600(±0.062) | 0.537(±0.015) |
| | | DenseNet | 0.624(±0.026) | 8.964E-5 | 0.661(±0.033) | 1.729E-3 | 0.668(±0.020) | 0.742(±0.093) | 0.454(±0.090) | 0.566(±0.016) | 0.657(±0.038) |
| | | ViT | 0.649(±0.020) | 6.302E-6 | 0.716(±0.024) | 4.420E-3 | 0.698(±0.024) | 0.600(±0.093) | 0.709(±0.084) | 0.670(±0.057) | 0.654(±0.045) |
| | | Swin Transformer | 0.717(±0.016) | 1.081E-4 | 0.757(±0.003) | 3.696E-4 | 0.742(±0.006) | 0.857(±0.067) | 0.563(±0.068) | 0.653(±0.035) | 0.813(±0.072) |
| Radiological report | our model | Chinese RoBERTa | **0.865(±0.012)** | **-** | **0.815(±0.011)** | **-** | **0.807(±0.012)** | **0.942(±0.019)** | 0.609(±0.036) | **0.697(±0.021)** | **0.917(±0.025)** |

| Data type | Model type | Model | Huashan Hospital, Fudan University (DH-IDH-1) | | | | | | | | |
|---|---|---|---|---|---|---|---|---|---|---|---|
| | | | AUC | *p* (AUC) | ACC | *p* (ACC) | F1-score | SEN | SPEC | PPV | NPV |
| Image | w/o segmentation | DeepRisk | 0.634(±0.078) | 1.188E-3 | 0.558(±0.031) | 7.363E-4 | 0.513(±0.030) | 0.420(±0.142) | 0.702(±0.132) | 0.337(±0.054) | 0.779(±0.019) |
| | | 3D MedMNIST | 0.617(±0.013) | 1.015E-4 | 0.610(±0.030) | 7.582E-4 | 0.605(±0.033) | 0.486(±0.176) | 0.644(±0.135) | 0.323(±0.033) | 0.788(±0.028) |
| | w/ segmentation | 2D MedMNIST | 0.583(±0.076) | 2.482E-3 | 0.548(±0.023) | 6.081E-4 | 0.498(±0.031) | 0.310(±0.096) | 0.770(±0.072) | 0.323(±0.039) | 0.763(±0.016) |
| | | DenseNet | 0.622(±0.025) | 1.938E-4 | 0.595(±0.018) | 6.429E-6 | 0.583(±0.024) | 0.449(±0.096) | 0.735(±0.077) | 0.376(±0.022) | 0.795(±0.012) |
| | | ViT | 0.675(±0.064) | 7.778E-3 | 0.655(±0.034) | 5.023E-3 | 0.652(±0.033) | 0.318(±0.097) | 0.854(±0.044) | 0.433(±0.063) | 0.783(±0.019) |
| | | Swin Transformer | 0.838(±0.033) | 5.647E-1 | 0.707(±0.037) | 7.995E-2 | 0.700(±0.038) | 0.388(±0.069) | 0.886(±0.027) | 0.546(±0.018) | 0.807(±0.013) |
| Radiological report | our model | Chinese RoBERTa | **0.848(±0.023)** | **-** | **0.772(±0.022)** | **-** | **0.765(±0.024)** | **0.535(±0.065)** | **0.912(±0.022)** | **0.684(±0.045)** | **0.850(±0.016)** |

Table S4: Detailed comparison between our language model, the image-based models, and three radiologists in terms of the AUC, ACC, and F1-score. The best result is highlighted in bold.

| Type | Model / Year | AUC | ACC | F1-score |
|---|---|---|---|---|
| Image-based models | DeepRisk | 0.575(±0.018) | 0.623(±0.025) | 0.630(±0.022) |
| | 3D MedMNIST | 0.622(±0.009) | 0.693(±0.010) | 0.679(±0.010) |
| | 2D MedMNIST | 0.711(±0.033) | 0.698(±0.024) | 0.704(±0.019) |
| | DenseNet | 0.748(±0.022) | 0.710(±0.013) | 0.715(±0.013) |
| | ViT | 0.689(±0.016) | 0.643(±0.028) | 0.655(±0.027) |
| | Swin Transformer | 0.692(±0.064) | 0.657(±0.023) | 0.667(±0.023) |
| Junior radiologists | 3 | - | 0.750 | 0.748 |
| | 3 | - | 0.789 | 0.778 |
| Experienced radiologist | 10 | - | **0.914** | **0.914** |
| Our report-based model | Chinese RoBERTa | **0.890(±0.009)** | 0.821(±0.021) | 0.818(±0.018) |

Table S5: Examples of the radiological reports from different hospitals. Note that the original radiological reports are in Chinese and the presented ones are translated versions. The image contrasts mentioned in each report are highlighted in bold.

| Hospital | Image contrasts | Relevant dataset | Radiological report (translated) |
|---|---|---|---|
| Beijing Tiantan Hospital | Full image contrasts | DT-IDH-1, DT-CI-1, DT-WHO-1, DT-BTC-1, | Localized brain tissue swelling is observed in the left frontal lobe, characterized by patchy and heterogeneous hyperintense **T1w** and **T2w** signals. The **FLAIR** image demonstrates heterogeneous hyperintense and hypointense signals. The **ADC** image reveals patchy restricted diffusion foci within the lesion, with slightly ill-defined boundaries, measuring approximately 42 × 38 × 28 mm in size. The adjacent brain tissue and sulci exhibit compression and deformation. The **T1c** image shows mild linear and patchy enhancement within the lesion. Scattered punctate regions with slightly higher **T2w** signals are noted in the left corona radiata and the left centrum semiovale, with indistinct boundaries. A small linear region exhibiting hyperintense **T1w** and **T2w** signals is identified in the inferior aspect of the right basal ganglia, characterized by relatively well-defined margins, suggestive of widened perivascular spaces. No abnormal signals are detected within the brain parenchyma. The size, position, and morphology of the ventricular system are within normal limits, with midline structures appropriately positioned, and the remaining sulci and fissures show no significant abnormalities. A small nodular abnormal signal is noted in the right maxillary sinus, associated with localized thickening of the sinus mucosa. The inner wall of the right orbit demonstrates irregularity. The size and position of the bilateral globes are within normal limits, with no significant abnormalities noted. |
| Beijing Tiantan Hospital | Lack of FLAIR images | DT-IDH-2, DT-CI-2, DT-WHO-2, DT-BTC-2, | A substantial mass exhibiting mixed signal characteristics, hyperintense on **T2w** image and isointense on **T1w** image, is identified in the right frontal and temporal regions. The mass has relatively well-defined margins with associated cystic changes, measuring approximately 47 × 48 × 53 mm. Surrounding the mass are extensive regions of hyperintense signals on both **T1w** and **T2w** imaging. On **T1c** image, the lesion shows marked enhancement. The **ADC** image reveals slightly reduced signal intensity within the lesion. The right lateral ventricle exhibits slight deformation, with midline structures generally positioned centrally. Multiple punctate foci of hyperintense signals on **T2w** and **T1w** images are noted in the pons, bilateral thalami, bilateral basal ganglia, left corona radiata, and left frontal lobe, which are characterized by indistinct margins. Sulci in the right hemisphere appear narrowed, whereas those in the left hemisphere are widened and deepened. A cerebrospinal fluid-like signal is observed within the sella turcica. |
| Beijing Tiantan Hospital | Lack of ADC images | DT-IDH-3 | Patchy areas of hyperintense signals on **T1w** and **T2w** images are identified in the left frontal, parietal, and temporal lobes, insula, basal ganglia, and thalamus, exhibiting mixed high signal intensity on **FLAIR** image with ill-defined margins. Mild punctate and linear enhancement is observed on **T1c** image, along with thickening and enhancement of the meninges in the surgical area. Scattered punctate low signal on **T2w** image is present in the left parietal cortex. The local sulci appear shallow, and the left lateral ventricle is enlarged and deformed, resulting in a mild rightward shift of midline structures. The **T1c** image shows multiple small ring-enhancing lesions with indistinct borders in the right corona radiata, bilateral basal ganglia, thalamus, and right temporal lobe. No significant abnormalities are noted in the paranasal sinuses, and both eyeballs appear normal in size and position. |
| Beijing Tiantan Hospital | Lack of both FLAIR and ADC images | DT-IDH-4 | Patchy hyperintense signals on **T1w** and high **T2w** images are noted in the cortex and subcortex of the right frontal lobe, characterized by heterogeneous signals and poorly defined margins. Small cystic regions are present within the lesion, which is surrounded by extensive edema with indistinct borders that affect the genu of the corpus callosum. Local sulci appear shallow or obliterated, and the frontal horn of the right lateral ventricle is compressed and distorted, accompanied by a leftward shift of midline structures.<br>There is no evidence of scalp soft tissue swelling. Bilateral mucosal thickening of the maxillary sinuses is observed, and both eyeballs exhibit normal size and position. The **T1c** image reveals the right frontal lesion with significant and heterogeneous enhancement, displaying an irregular morphology and well-defined borders, measuring approximately 45 × 46 × 38 mm. |
| The First Affiliated Hospital of Xinjiang Medical University | Full image contrasts | DX-IDH-1 | Irregular, mixed lesions with slightly hyperintense signals on **T1w** and **T2w** images are identified in the bilateral frontal cingulate gyri, the midline region of the brain, and the left superior frontal gyrus. The margins of the lesions display patchy low signal intensity on **T1w** image and high signal intensity on **T2w** image. Within the lesions, linear low signal intensity on **T2w** image is evident. On the **FLAIR** image, the lesions appear slightly hyperintense. In **ADC** image, the solid components of the lesions show slightly hyperintense signals. Extensive surrounding edema is noted. The **T1c** image reveals significant heterogeneous enhancement of the lesions, measuring approximately 7.88 cm × 5.75 cm. The body of the corpus callosum is involved and displaced inferiorly, resulting in compression and distortion of the frontal horns of the bilateral lateral ventricles, with local midline structures shifted to the right. |
| Huashan Hospital, Fudan University | Lack of T2w images | DH-IDH-1 | A mass located in the right frontal lobe is identified, displaying low signal intensity on **T1w** image and high signal intensity on **FLAIR** image, accompanied by extensive surrounding edema. The lesion demonstrates high signal intensity on **ADC** and has ill-defined margins. The **T1c** image reveals heterogeneous enhancement, with involvement of the body of the corpus callosum. Additionally, the right lateral ventricle is compressed and narrowed, resulting in a marked leftward displacement of midline structures. |

[10] Zhilu Zhang and Mert Sabuncu. Generalized cross entropy loss for training deep neural networks with noisy labels. *Advances in neural information processing systems*, 31, 2018.
